# Differential Levodopa Responsiveness and Motor Complication Trajectories in Parkinson’s Disease by α-Synuclein Seed Amplification Assay Status

**DOI:** 10.64898/2026.04.21.26350973

**Authors:** Houman Azizi, Seyed-Mohammad Fereshtehnejad, Roqaie Moqadam, Mahsa Dadar, Andrew Siderowf, Alain Dagher, Yashar Zeighami

## Abstract

**Background:** Cerebrospinal fluid (CSF) α-synuclein seed amplification assay (SAA) has emerged as a diagnostic biomarker for Parkinson’s disease (PD) and has been linked to differences in disease severity and progression. However, whether SAA status predicts responsiveness to levodopa remains unknown. We investigated the longitudinal association between SAA status, levodopa responsiveness, dopaminergic denervation, and motor complications in sporadic PD.

**Methods:** In this longitudinal analysis, PD participants from the Parkinson’s Progression Markers Initiative (PPMI) cohort with CSF SAA testing who initiated levodopa treatment were included. SAA^-^ and SAA^+^ patients were matched on sex, age, and disease duration at treatment initiation. Motor severity was assessed using MDS-UPDRS Part III, with proportional and absolute responsiveness derived from ON and OFF medication states. Motor complications were assessed using MDS-UPDRS Part IV, and dopaminergic dysfunction was quantified using caudate DAT-SPECT. Linear mixed-effects models examined longitudinal differences as a function of SAA status.

**Findings:** In this analysis, 40 SAA^-^ patients were compared to 183 matched SAA^+^ patients. SAA^+^ patients showed a slower rate of ON-state motor progression than SAA^-^ patients (0.87 vs 3.47 points/year; p = 0.01). Consistently, proportional levodopa responsiveness increased over time in SAA^+^ patients while declining in SAA^-^ patients (p = 0.036). These differences were accompanied by lower caudate DAT binding at treatment initiation in SAA^-^ patients (p = 0.002) and faster dopaminergic decline over time (p = 0.008). Although SAA^+^ patients had fewer motor complications at treatment initiation, their progression was similar.

**Interpretation:** CSF α-synuclein SAA status is associated with divergent levodopa response in PD, with SAA^+^ patients showing sustained and progressively greater motor benefit, while SAA^-^ patients show declining responsiveness. Faster dopaminergic denervation in SAA^-^ patients may underlie this difference. SAA status captures clinically relevant heterogeneity that may inform patient stratification and therapeutic decision-making.

## Introduction

Aggregation of misfolded alpha-synuclein (α-Syn) within Lewy bodies plays a key role in pathology of Lewy body disorders such as Parkinson’s disease (PD)^1,2^. The cerebrospinal fluid (CSF) α-synuclein seed amplification assay (SAA) has emerged as a highly sensitive and specific biomarker^3^ for differentiating PD from healthy controls using both clinical diagnosis^4,5^ and postmortem confirmation^6,7^. Beyond its diagnostic utility, CSF SAA status has shown clinical associations with disease severity and progression in Lewy body diseases^8^. SAA status in participants with dementia with Lewy bodies has been associated with differences in postural instability and motor severity, cognitive performance, and Hoehn & Yahr clinical staging^9^. In PD, SAA status has further been linked to differences in age of disease onset as well as the rate of progression along clinical milestones including postural instability^10^. Despite some reported inconsistencies, quantitative kinetic parameters of SAA have been shown to correlate with Movement Disorder Society Unified Parkinson’s Disease Rating Scale Part III (MDS-UPDRS Part III)^11^ and Hoehn & Yahr stage^12^, and have been linked to future cognitive decline and likelihood of presenting as diffuse malignant subtype in PD^13,14^.

Despite these advances, the relationship between SAA status and motor progression remains inconsistent, reflecting substantial clinical heterogeneity. Some studies have reported significant differences in MDS-UPDRS Part III scores between SAA positive (SAA^+^) and negative (SAA^-^) patients, while others have reported comparable motor severity^9,10^. This relationship also varies among different PD subgroups^4^. Furthermore, PD patients show considerable variability in levodopa responsiveness and motor improvement rates among clinically comparable individuals^15,16^, and it remains unknown how the subgroup of individuals with SAA^-^ PD clinically respond to levodopa therapy. Another potential factor contributing to this variability is the degree of striatal dopaminergic transporter availability as measured by dopamine transporter single-photon emission computed tomography (DAT-SPECT). Striatal DAT binding correlates with motor symptom burden and declines with disease progression in PD^17,18^, and has further been linked to CSF α-synuclein levels^19,20^.

Sex differences exist in PD, with males having up to 1.4 times the risk of developing this disease^21^ and present with more severe motor impairment and faster disease progression^22^, while females tend to have a later age of onset, and a tremor-dominant phenotype^23^. Sex differences are also seen in treatment responses where females show greater levodopa bioavailability^24,25^ and a higher susceptibility to levodopa-induced dyskinesias and wearing-off effects^26,27^. However, whether sex interacts with SAA status to influence levodopa responsiveness remains unclear. Given that levodopa is the gold standard treatment for motor symptoms in PD, understanding whether α-synuclein pathology influences medication responsiveness represents a critical knowledge gap.

In this study, we took advantage of the longitudinal assessments available from the Parkinson’s Progression Markers Initiative (PPMI) to investigate whether CSF SAA status predicts levodopa responsiveness in PD. Understanding whether α-synuclein pathology, as detected by the SAA, predicts dopaminergic treatment efficacy could enable biomarker-based stratification in PD, ultimately supporting more targeted treatment planning and clinical trial design.

## Methods

### Participants

Data were obtained from the Parkinson’s Progression Markers Initiative (PPMI), a multicenter longitudinal prospective cohort^28^. Participants enrollment criteria included a clinical diagnosis of PD, time since diagnosis of less than two years at enrollment, and abnormal dopamine transporter single-photon emission computed tomography (DAT-SPECT) imaging. Participants underwent CSF SAA testing at their baseline visit, as previously described^4,29^. We restricted our sample to participants with sporadic PD (sPD) diagnosis who had undergone CSF SAA testing and initiated levodopa medication. Individuals with genetic forms of PD such as LRRK2- or SNCA-associated disease as well as participants with inconclusive or type II SAA results were excluded. Additionally, participants with levodopa equivalent daily dose (LEDD) values exceeding three standard deviations from the sample mean were excluded, and analyses were restricted to visits from treatment initiation up to 8 years post-treatment initiation in which participants had MDS-UPDRS Part III scores recorded in both ON and OFF medication states.

### Matching

Given the demographic differences between SAA^+^ and SAA^-^ groups, we performed matching based on sex, age, and time since clinical diagnosis at the first post-medication visit to ensure the findings are not impacted by these differences. Matching was performed separately for the main and sex stratified analysis to maximize the sample size. For continuous variables (age and time since diagnosis), a tolerance of 10% deviation from the sample mean was used. Variable-ratio nearest-neighbor matching^30^ was applied, matching each SAA^-^ participant to up to 5 SAA^+^ participants. When fewer suitable matches were available, smaller ratios were retained to maximize the number of included participants while preserving covariate balance. In total, 40 SAA^-^ patients were matched to 183 SAA^+^ patients at an approximate 1:4 ratio.

### Outcome Measures

Motor severity was assessed using MDS-UPDRS Part III in both ON and OFF states at each visit. Medication responsiveness was quantified as: (1) Proportional medication responsiveness, defined as the change in motor scores from the OFF to the ON state relative to the OFF state score (i.e., [UPDRS-III_OFF_-UPDRS-III_ON_]/UPDRS-III_OFF_); (2) Absolute medication responsiveness, defined as the difference between OFF and ON state scores^31^.

Motor complications were assessed using MDS-UPDRS Part IV total score, as well as three subcategories: (1) dyskinesia: sum of items 4.1-4.2 (time spent with dyskinesias and functional impact of dyskinesias), (2) motor fluctuations: sum of items 4.3-4.5 (time spent in the OFF state, functional impact of fluctuations, complexity of motor fluctuations), and (3) painful OFF-state dystonia: item 4.6^11^.

### Dopamine Transporter Imaging

DAT-SPECT imaging from post-treatment visits were used to assess dopaminergic dysfunction in SAA^−^ and SAA^+^ groups, using the mean caudate nucleus binding ratio (SBR), calculated as previously described^32^. Among our matched sample, 29 SAA^−^ and 136 SAA^+^ patients had DAT-SPECT imaging available within 2 years of treatment initiation. For longitudinal analysis, only visits with both DAT-SPECT and MDS-UPDRS Part III available were included, with follow-up was restricted to 4 years due to limited data availability beyond this timepoint (N = 25 SAA^-^, 129 SAA^+^).

### Statistical Analysis

All analyses were performed using linear mixed-effects models. Time (years from treatment initiation), SAA, and medication status are the primary fixed effect variables. In all models described below, age at treatment initiation, sex, and LEDD were included as covariates. Models also included random intercept and slope (for time variable) per participant. The full specification of the models are provided in the Supplementary Methods. Statistical significance was set at α = 0.05.

#### Motor progression as a function of medication state and SAA status

To characterize how motor progression trajectories differed between SAA^+^ and SAA^-^ groups across medication states, we modeled MDS-UPDRS Part III scores by treating ON and OFF motor scores from each visit as separate observations per participant. We used a three-way interaction between time, SAA status, and medication state (OFF vs. ON), allowing us to examine whether the rate of motor progression differed between SAA groups in a medication state–dependent manner.

#### Dopamine transporter availability

We compared caudate DAT binding ratios at the first post-medication visit between SAA groups using a two-sided Wilcoxon rank-sum test. Longitudinal caudate DAT binding was modeled as a function of time, SAA status, and their interaction. Due to limited longitudinal DAT-SPECT data availability, only a random intercept was included in this model.

#### Medication responsiveness

Proportional and absolute medication responsiveness (as previously defined) were modeled separately as a function of time, SAA status, and their interaction, to examine differences at the treatment initiation and over time.

#### Wilcoxon rank-sum tests for individual intercepts and slopes

In addition to the above-mentioned mixed-effects models, we derived individual-level estimates of baseline values (intercepts) and annual rates of change (slopes) for each participant by combining the fixed-effect estimates with each participant’s random effects from each model. These values were compared between SAA groups using two-sided Wilcoxon rank-sum tests to assess group differences at the treatment initiation and their rate of change over time.

#### Motor complications

We separately modelled the MDS-UPDRS Part IV total score as well as the dyskinesia, the motor fluctuations, and the painful OFF-state dystonia subscores using an interaction between time and SAA status.

## Results

Our sample included 40 SAA^−^ and 183 SAA^+^ patients with sPD. At treatment initiation, SAA^−^ and SAA^+^ patients were matched based on age (67.8 vs. 66.6 years, respectively; p = 0.38), sex (40.0% vs. 37.2% female, respectively; p = 0.88), and disease duration (1.43 vs. 1.37 years, respectively; p = 0.69). The mean post treatment follow-up duration is 2.3 ± 2.0 for SAA^−^ and 3.3 ± 2.4 years for SAA^+^ patients (p = 0.01) (4.0 and 5.2 years from enrollment). Complete participant characteristics at the first post treatment visit are presented in Table 1.

**Table 1.** Group characteristics at first ON-dopaminergic medication visit.

| Measure | SAA Negative | SAA Positive | Test Statistic | P-Value |
| --- | --- | --- | --- | --- |
| **N** | 40 | 183 |  |  |
| **N Visits, total (SD)** | 8.40 (5.25) | 10.45 (6.82) | **t(71.0) = -2.11** | **0.038** |
| **N Visits Included in Analysis (SD)** | 2.83 (1.53) | 3.61 (2.04) | **t(72.7) = -2.76** | **0.007** |
| **Age at Treatment Initiation, years (SD)** | 67.82 (8.22) | 66.57 (7.12) | t(52.6) = 0.89 | 0.375 |
| **Age at Baseline Visit, years (SD)** | 67.12 (8.20) | 65.92 (7.09) | t(52.5) = 0.86 | 0.395 |
| **Sex, Female (%)** | 16 (40.0%) | 68 (37.2%) | X<sup>2</sup>(1) = 0.02 | 0.876 |
| **Total Duration in the Study, years (SD)** | 3.96 (3.06) | 5.16 (3.98) | **t(71.0) = -2.11** | **0.039** |
| **Follow-Up Since Medication Initiation, years (SD)** | 2.34 (1.98) | 3.28 (2.42) | **t(67.2) = -2.61** | **0.011** |
| **LEDD, mg/day (SD)** | 295.31 (177.52) | 288.42 (198.39) | t(62.2) = 0.22 | 0.828 |
| **Hoehn & Yahr Stage (SD)** | 1.82 (0.50) | 1.81 (0.47) | t(55.1) = 0.19 | 0.852 |
| **Schwab & England ADL, % (SD)** | 90.00 (7.07) | 89.92 (8.57) | t(66.6) = 0.06 | 0.949 |
| **Education, years (SD)** | 16.29 (3.28) | 15.73 (3.30) | t(48.4) = 0.92 | 0.361 |
| **MoCA Score, adjusted (SD)** | 26.45 (3.12) | 26.63 (2.87) | t(54.8) = -0.33 | 0.74 |
| **Geriatric Depression Scale (SD)** | 2.25 (2.17) | 2.65 (2.93) | t(74.7) = -0.97 | 0.334 |
| **STAI Score, total (SD)** | 59.25 (13.07) | 64.99 (20.56) | **t(88.6) = -2.22** | **0.029** |
| **Apathy Score (SD)** | 0.35 (0.62) | 0.37 (0.70) | t(62.3) = -0.15 | 0.885 |
| **QUIP Score (SD)** | 0.25 (0.63) | 0.24 (0.65) | t(59.1) = 0.08 | 0.94 |
| **SCOPA-AUT, Autonomic (SD)** | 12.12 (6.61) | 11.97 (7.10) | t(61.1) = 0.13 | 0.896 |
| **RBD Questionnaire Score (SD)** | 2.88 (2.75) | 4.13 (3.15) | **t(64.2) = -2.54** | **0.014** |
| **UPSIT Percentile at Closest Visit (SD)** | 49.95 (28.07) | 13.60 (14.58) | **t(41.4) = 7.76** | **< 0.001\*** |
| **MDS-UPDRS I (SD)** | 7.00 (4.30) | 8.03 (8.56) | t(117.0) = -1.11 | 0.271 |
| **MDS-UPDRS II (SD)** | 9.85 (4.75) | 8.45 (5.74) | t(66.3) = 1.62 | 0.11 |
| **MDS-UPDRS III, OFF Medication (SD)** | 27.89 (9.54) | 26.68 (12.25) | t(66.6) = 0.67 | 0.503 |
| **MDS-UPDRS III, ON Medication (SD)** | 21.56 (9.52) | 20.15 (9.64) | $t(56.5) = 0.84$ | 0.405 |
| **Tremor Score, OFF Medication (SD)** | 0.75 (0.46) | 0.92 (0.56) | $t(63.8) = -1.97$ | 0.054 |
| **PIGD Score, OFF Medication (SD)** | 0.50 (0.38) | 0.34 (0.31) | **$t(48.8) = 2.44$** | **0.018** |
| **MDS-UPDRS IV** |  |  |  |  |
| **Total (SD)** | 0.78 (2.03) | 0.44 (1.25) | $t(45.7) = 0.99$ | 0.325 |
| **Dyskinesia Subscore (SD)** | 0.02 (0.16) | 0.04 (0.32) | $t(120.0) = -0.38$ | 0.701 |
| **Motor Fluctuations Subscore (SD)** | 0.65 (1.69) | 0.37 (1.09) | $t(46.4) = 1.02$ | 0.314 |
| **Painful OFF-State Dystonia (SD)** | 0.10 (0.63) | 0.04 (0.24) | $t(41.5) = 0.61$ | 0.547 |
| **DATScan Caudate, SBR (SD)** | 0.54 (0.23) | 0.75 (0.27) | **$t(46.0) = -4.27$** | **< 0.001\*** |
| **DATScan Putamen, SBR (SD)** | 0.56 (0.23) | 0.64 (0.19) | $t(36.3) = -1.74$ | 0.09 |

Demographic, clinical, neuropsychiatric, and imaging characteristics of α-Syn SAA<sup>+</sup> and SAA<sup>-</sup> SPD patients at their first ON-dopaminergic medication visit within two years of treatment initiation. For each measure, values represent the group mean (SD) for continuous variables or count (percentage) for categorical variables, derived from the first available post-medication visit per patient at which the measure was recorded (within the first 2 years of treatment initiation). Between-group comparisons were performed using Welch's two-sample t-test for continuous variables and Pearson's chi-squared test for categorical variables. P-values marked with an asterisk remained significant after False Discovery Rate (FDR) correction. LEDD = levodopa equivalent daily dose; HY = Hoehn & Yahr disease stage; ADL = activities of daily living; MoCA = Montreal Cognitive Assessment; STAI = State-Trait Anxiety Inventory; QUIP = Questionnaire for Impulsive-Compulsive Disorders in Parkinson's Disease; SCOPA-AUT = Scales for Outcomes in Parkinson's Disease – Autonomic; RBD = REM sleep behavior disorder; UPSIT = University of Pennsylvania Smell Identification Test; MDS-UPDRS = Movement Disorder Society Unified Parkinson's Disease Rating Scale; PIGD = postural instability and gait difficulty; DATScan SBR = dopamine transporter scan striatal binding ratio; SAA = seed amplification assay; αSyn = α-synuclein.

We examined the effect of levodopa on motor progression by SAA status using a three way interaction between time, medication state, and CSF SAA status. This interaction was significant (β = -1.31, p = 0.01; Figure 1). In the ON medication state, SAA^+^ patients progressed at a significantly slower rate compared to SAA^−^ patients (β = -2.61, p < 0.001), while in the OFF medication state, SAA^−^ patients showed a trend towards faster deterioration compared to SAA^+^ patients (Time and SAA interaction: β = -1.29, p = 0.066). Post hoc analyses showed that levodopa significantly reduced the rate of motor progression by 49.3% in SAA^+^ patients (from 1.86 to 0.94 points annual progression; p < 0.001). In the SAA^-^ patients, while the medication reduced the motor symptoms, the rate of progression remained similar between OFF and ON states (p = 0.36).

**Figure 1:**
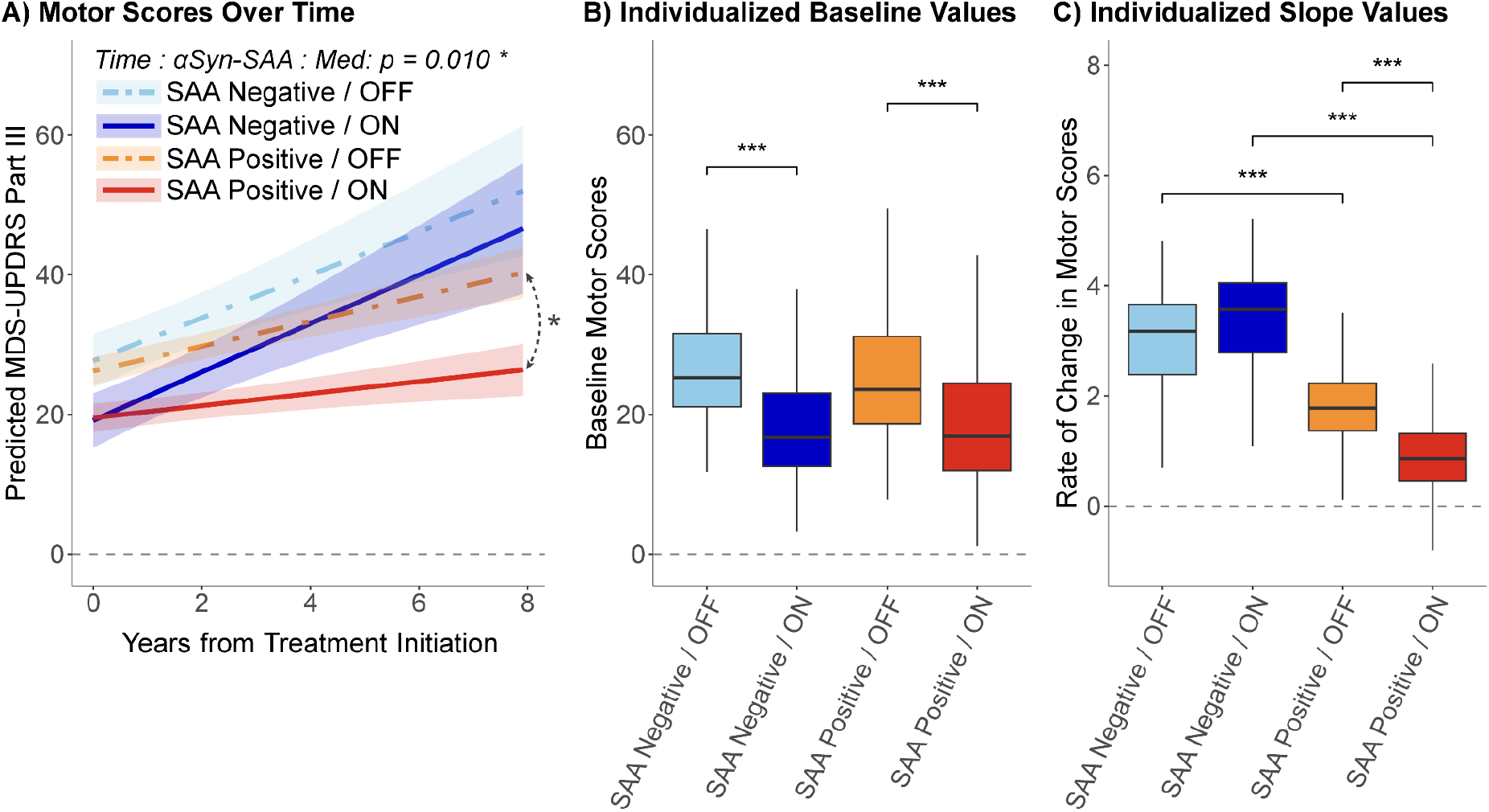
UPDRS III motor score progression by SAA status and medication state. **(A)** Longitudinal predicted change in MDS-UPDRS Part III motor score for α-Syn SAA^+^ (red) and SAA^−^ (blue) sPD patients based on ON (solid lines, darker colors) and OFF (dotted lines, lighter colors) medication state. Curved arrows with asterisks show statistically significant slope differences. **(B)** Individual subject motor scores at treatment initiation, calculated as the sum of fixed and random intercepts derived from the mixed-effects model. **(C)** Individual subject annual rates of motor score change, calculated as the sum of fixed and random slopes derived from the mixed-effects model. In both (B) and (C), statistically significant differences between groups assessed by Wilcoxon rank-sum test are indicated by asterisks (ns non-significant, * p < 0.05, ** p < 0.01, *** p < 0.001). αSyn = α-synuclein; SAA = seed amplification assay; MDS-UPDRS = Movement Disorder Society Unified Parkinson’s Disease Rating Scale; Med = Medication State (ON or OFF).

To examine whether the observed differences in Levodopa responsiveness between SAA groups were accompanied by differences in presynaptic dopaminergic transporter availability, we compared dopamine transporter (DAT) binding ratios obtained from DAT-SPECT imaging in the caudate nucleus. In the matched sample, 29 SAA^−^ and 136 SAA^+^ patients had DAT-SPECT imaging available within 2 years (average 0.56 years) of treatment initiation. At the first post-medication visit, SAA^+^ patients showed significantly higher caudate DAT binding compared to SAA^−^ patients (mean SBR: 0.75 ± 0.27 vs. 0.54 ± 0.23, respectively; p < 0.001; Figure 2A).

**Figure 2:**
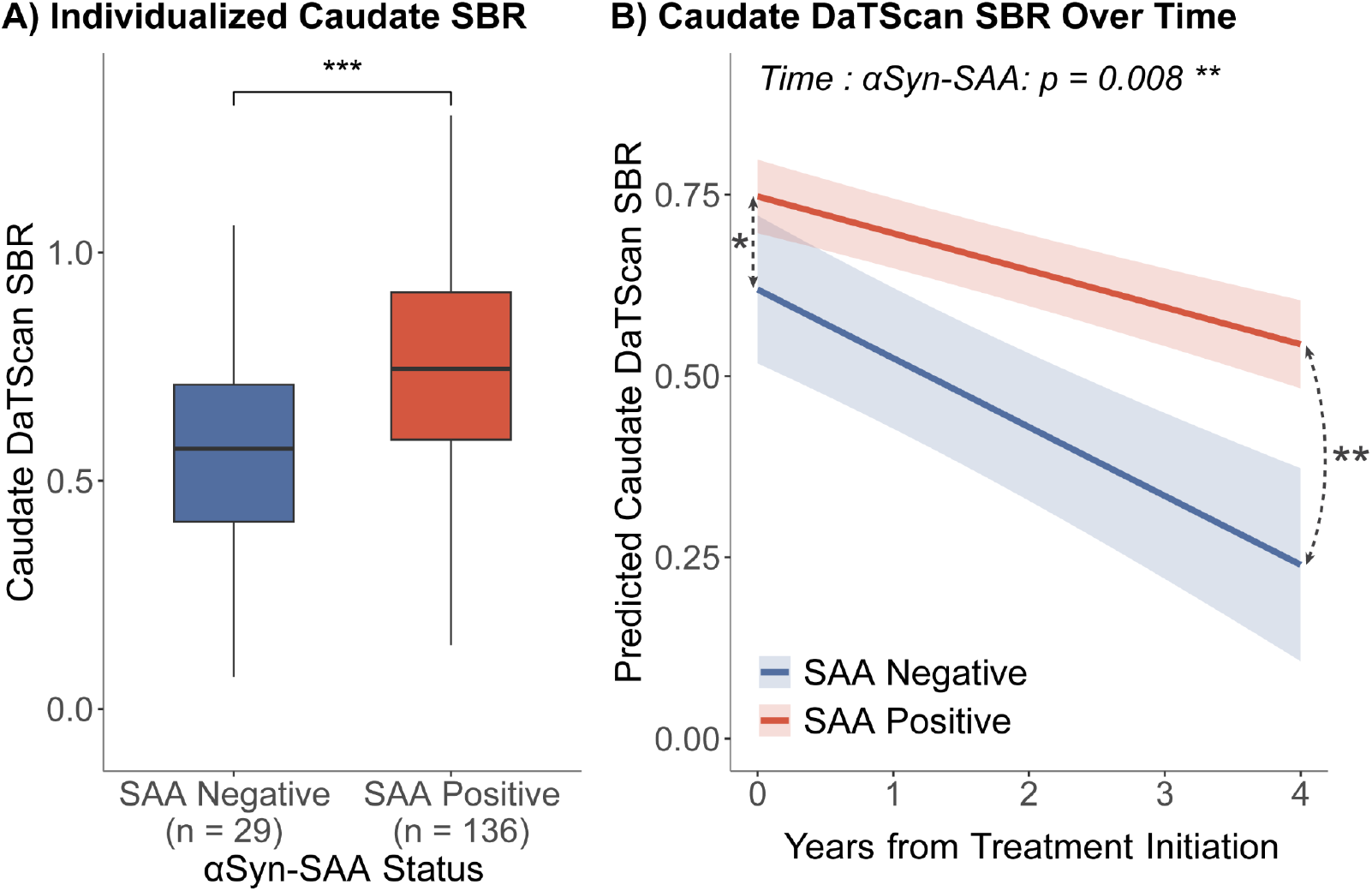
Caudate DaTScan SBR by SAA status. **(A)** Raw individual subject DaT Scan SBR at earliest visit within two years of medication initiation. Statistically significant differences between groups as assessed by Wilcoxon rank-sum test are indicated by asterisks (ns non-significant, * p < 0.05, ** p < 0.01, *** p < 0.001) **(B)** Longitudinal predicted change in Caudate DaT Scan SBR for α-Syn SAA^+^ (red) and SAA^−^ (blue) sPD patients. Curved arrows with asterisks show statistically significant slope differences. Straight arrows with asterisks show statistically significant intercept differences. αSyn = α-synuclein; SAA = seed amplification assay; DaTScan = dopamine transporter scan; SBR = striatal binding ratio.

We further examined whether caudate DAT binding trajectories differed between SAA groups over time. Both groups showed a decline in caudate DAT binding over time (β = -0.095, p < 0.001), however the rate of decline was significantly slower in SAA^+^ compared to SAA^−^ patients (Time and SAA interaction: β = 0.044, p = 0.008; Figure 2B). These results suggest that SAA^−^ patients not only have lower caudate dopamine transporter availability at treatment initiation but also experience a faster rate of dopaminergic denervation over time, providing a potential mechanism for progressively diminishing medication responsiveness observed in this group.

We examined whether medication responsiveness differed by SAA status using proportional and absolute mediation responsiveness in SAA^−^ and SAA^+^ patients. Proportional medication responsiveness, [(UPDRS-III_OFF_-UPDRS-III_ON_)/UPDRS-III_OFF_], was comparable at treatment initiation (β = -0.024, p = 0.47; 28.0% vs 29.9% in SAA^+^ and SAA^−^, respectively). Over time, proportional Levodopa responsiveness significantly diverged between groups as SAA^+^ patients showed 1.2% greater medication responsiveness per year (Time and SAA interaction: β = 0.028, p = 0.036) while medication responsiveness decreased 1.7% per year in SAA^−^ patients (β = -0.017, p = 0.20), indicating persistently higher responsiveness in SAA^+^ patients throughout the follow-up period (Figure 3).

**Figure 3:**
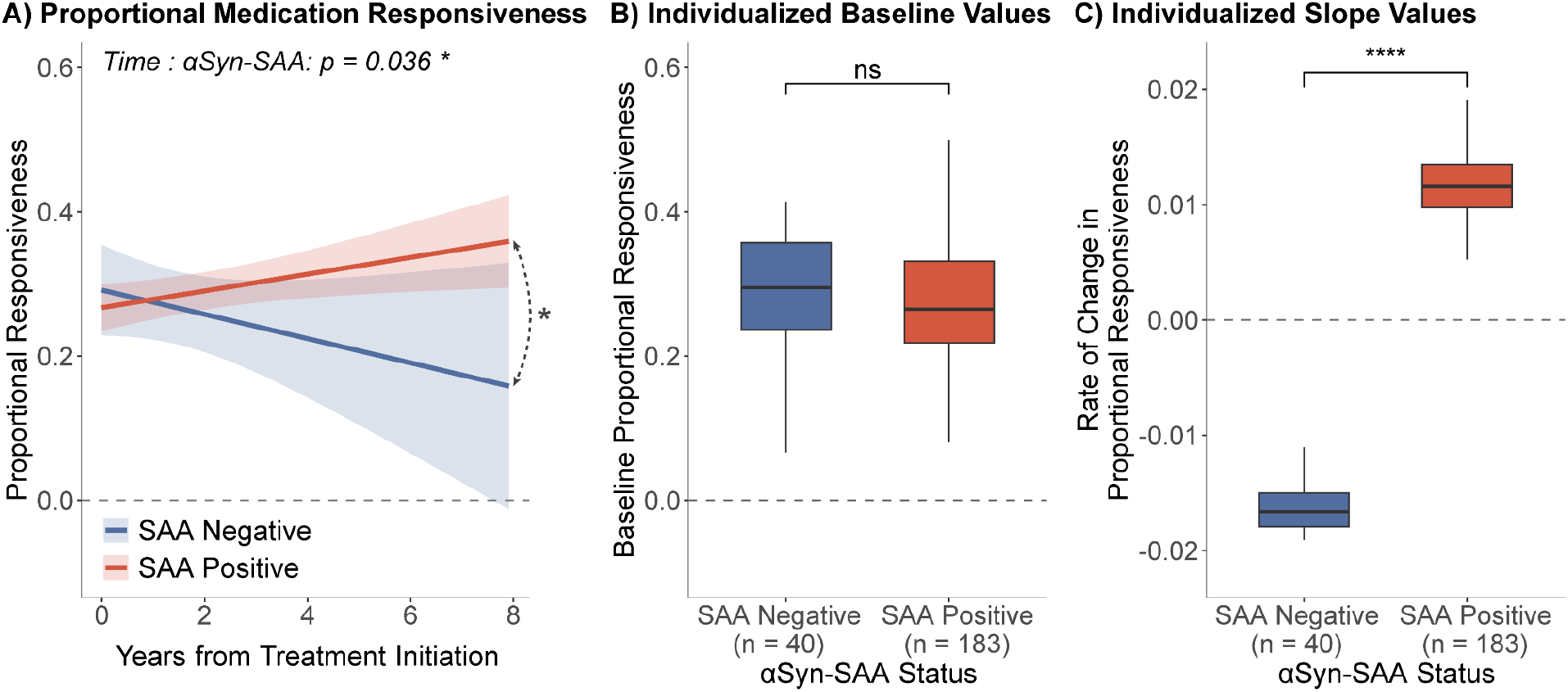
Longitudinal change in proportional medication responsiveness by SAA status. **(A)** Longitudinal predicted change in proportional medication responsiveness, calculated as MDS-UPDRS-III_OFF_ - UPDRS-III_ON_] / MDS-UPDRS-III_OFF_, for α-Syn SAA^+^ (red) and SAA^−^ (blue) sPD patients over years from treatment initiation. Curved arrows with asterisks show statistically significant slope differences. **(B)** Individual subject proportional medication responsiveness at treatment initiation, calculated as the sum of fixed and random intercepts derived from the mixed-effects model. **(C)** Individual subject annual rates of change in proportional medication responsiveness, calculated as the sum of fixed and random slopes derived from the mixed-effects model. In both (B) and (C), statistically significant differences between groups assessed by Wilcoxon rank-sum test are indicated by asterisks (ns non-significant, * p < 0.05, ** p < 0.01, *** p < 0.001). αSyn = α-synuclein; SAA = seed amplification assay; MDS-UPDRS = Movement Disorder Society Unified Parkinson’s Disease Rating Scale.

For absolute responsiveness, (UPDRS-III_OFF_ - UPDRS-III_ON_), at the start of the treatment, SAA^+^ and SAA^−^ patients did not show a statistically significant difference (β = -1.41, p = 0.16; 6.6 and 7.8 points reduction, respectively). However, over time, the medication responsiveness deviated between groups with a 1.05 points/year increase in SAA^+^ patients while remaining relatively stable in SAA^−^ patients (0.17 points/year increase), although this difference did not reach statistical significance (Time and SAA interaction β = 0.89, p = 0.098; Figure 4).

**Figure 4:**
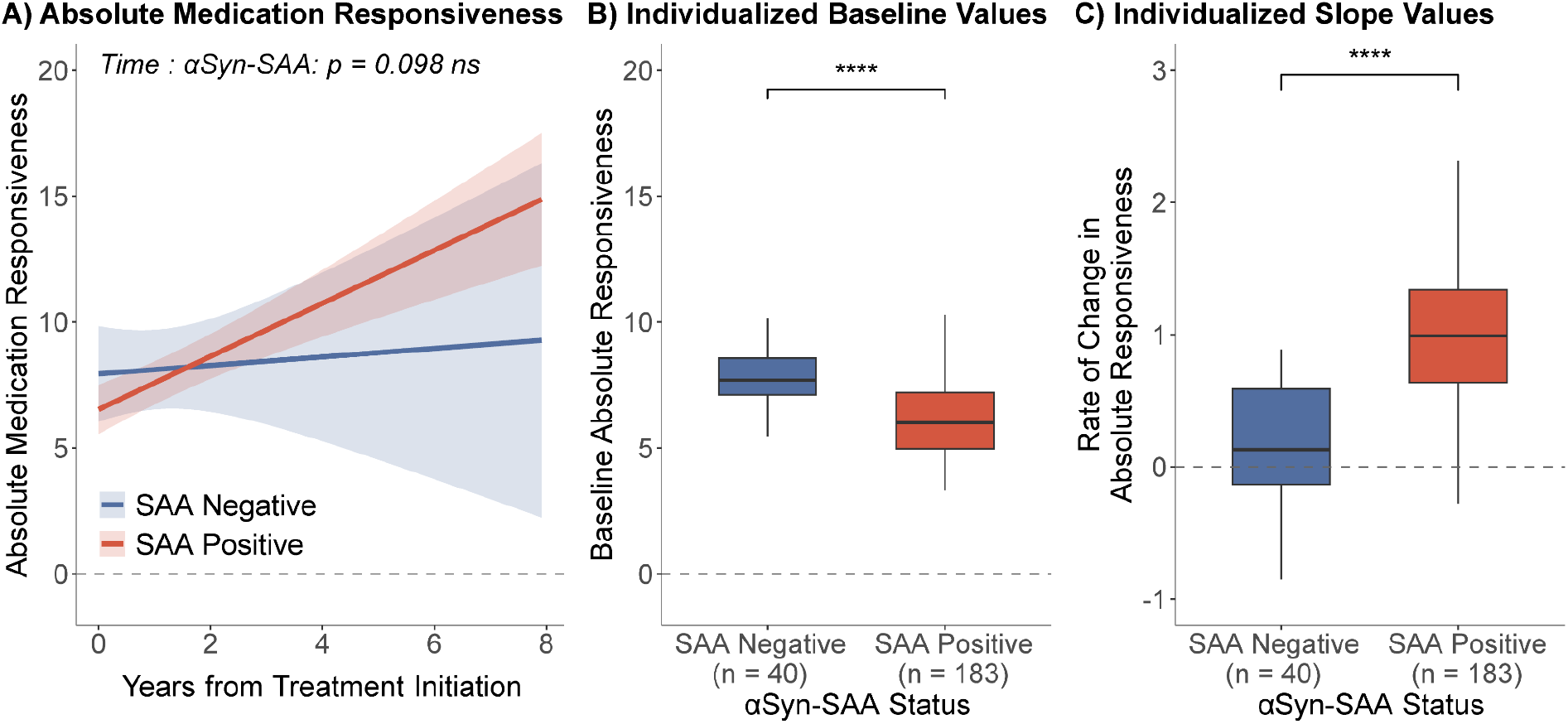
Absolute medication responsiveness by SAA status. **(A)** Longitudinal predicted change in absolute medication responsiveness, calculated as MD-UPDRS-III_OFF_ - MDS-UPDRS-III_ON_, for α-Syn SAA^+^ (red) and SAA^−^ (blue) sPD patients over years from treatment initiation. **(B)** Individual subject absolute medication responsiveness at treatment initiation, calculated as the sum of fixed and random intercepts derived from the mixed-effects model. **(C)** Individual subject annual rates of change in absolute medication responsiveness, calculated as the sum of fixed and random slopes derived from the mixed-effects model. In both (B) and (C), statistically significant differences between groups assessed by Wilcoxon rank-sum test are indicated by asterisks (ns non-significant, * p < 0.05, ** p < 0.01, *** p < 0.001). αSyn = α-synuclein; SAA = seed amplification assay; MDS-UPDRS = Movement Disorder Society Unified Parkinson’s Disease Rating Scale.

We next examined motor complications (MDS-UPDRS Part IV) over time between SAA^+^ and SAA^−^ patients. At treatment initiation, SAA^+^ patients had lower total scores (β = -1.07, p = 0.001) and motor fluctuation scores (β = −0.90, p < 0.001) compared to SAA^-^ patients (Supplementary Figure S1). However, the rate of progression remained comparable between SAA groups across all motor complication categories (all time and SAA interaction p-values > 0.15). These findings indicate that while SAA^+^ patients present with lower motor complications at treatment initiation, their rate of progression of these complications is similar to SAA^−^ patients.

We repeated all analyses separately in males and females to assess sex-specific effects. In the OFF medication state, SAA^+^ and SAA^−^ females showed comparable motor progression (Time and SAA interaction: p = 0.49), while SAA^+^ males showed a trend toward slower progression compared to SAA^−^ males (Time and SAA interaction: β = -1.67, p = 0.07). The three-way interaction between time, SAA status, and medication state showed a similar direction of effect in both sexes, with larger levodopa-induced motor improvement in SAA^+^ patients, though these effects only showed trends given the smaller sample sizes (females: β = -1.73, p = 0.053; males: β = -1.11, p = 0.074; Supplementary Figure S2).

The proportional medication response did not differ based on SAA status at treatment initiation in either sex (males: p = 0.74; females: p = 0.14). Over time, SAA^−^ females showed a significantly lower proportional responsiveness than SAA^+^ females (Time SAA interaction: p = 0.044; Supplementary Figure S3), while this difference was not observed in males (p = 0.23). SAA^+^ and SAA^−^ patients further showed comparable absolute medication responsiveness at treatment initiation (males: p = 0.54; females p = 0.19). SAA^+^ patients showed larger improvements in absolute medication responsiveness compared to SAA^−^ patients in both sexes. While this difference was larger in females, it did not reach statistical significance in either sex (Time SAA interaction: p = 0.078 in females; p = 0.60 in males).

Sex stratified analyses of motor complications were consistent with the combined group analysis, with no significant progression rate differences in either sex. Consistent with the combined analysis, SAA^+^ patients showed lower total motor complication scores (males: β = -1.09, p = 0.006; females: β = -1.23, p = 0.035) and motor fluctuation scores (males: β = -0.98, p = 0.009; females: β = -0.84, p = 0.03) at treatment initiation compared to SAA^−^ patients in both sexes. Together, these findings suggest that while the directions of effects were broadly consistent, the differential levodopa benefit on motor progression was more pronounced in females.

## Discussion

In this longitudinal study of sPD patients’ response to dopaminergic therapy, we observed differential response based on CSF α-synuclein SAA status. SAA^+^ patients had a significantly slower rate of motor progression in the ON medication state compared to SAA^−^ patients, indicating a treatment-dependent divergence whereby SAA^+^ patients show a sustained benefit from levodopa over the course of the disease. In contrast, motor progression in the OFF state was largely comparable between groups, while SAA^−^ patients showed a trend toward more aggressive OFF-state progression, consistent with prior reports of faster clinical progression and more pronounced imaging abnormalities in SAA^−^ PD patients even at early drug-naive stage^10^. Together, these findings suggest that SAA status is associated with differential longitudinal responsiveness to levodopa, rather than substantial differences in baseline disease severity.

The observed differences in medication responsiveness provides important insight into the role of α-synuclein pathology in sPD and were further accompanied by differences in presynaptic dopamine transporter availability measured by DAT-SPECT. SAA^−^ patients showed significantly lower initial striatal binding ratio in the caudate and a faster rate of dopaminergic denervation over time. These findings provide a potential neurobiological mechanism for the progressively diminishing levodopa responsiveness in the SAA^−^ group, whereby the capacity of converting exogenous levodopa into dopamine is progressively reduced due to presynaptic dopamine terminal loss^33,34^. Consistent with prior evidence linking caudate dopaminergic depletion to higher levodopa dose requirements in early PD^35^, the observed higher preservation of dopamine transporter in SAA^+^ patients supports the more sustained pharmacological response seen in this group. These findings along with the marginally faster OFF-state motor progression suggest that SAA^−^ sPD patients may undergo a more aggressive pattern of nigrostriatal degeneration that compromises both motor functions as well as the capacity to benefit from dopaminergic therapy.

Despite similar OFF-state medication responsiveness between groups at treatment initiation, SAA^−^ patients demonstrated a progressive decline in proportional responsiveness over time, whereas SAA^+^ patients showed an increase in proportional improvement and a greater increase in absolute medication responsiveness over time. By explicitly modeling longitudinal trajectories and separating ON and OFF medication states, we showed that differences between SAA groups emerge primarily in the context of treatment rather than baseline severity. This suggests that synucleinopathy might impact the variability in treatment response rather than explaining cross-sectional differences in motor impairment. While the response rate was consistently lower in the SAA^−^ individuals, they nevertheless demonstrated a measurable and sustained motor response to levodopa, with an improvement of around 30%, consistent with a positive levodopa response early in the disease course. Our findings suggest that α-synuclein pathology captures biologically meaningful heterogeneity within clinically diagnosed PD that is relevant to dopaminergic treatment responsiveness.

Compared with SAA^+^ patients, individuals with SAA^−^ sPD had more motor complications at treatment initiation. Despite this initial difference, they showed a comparable rate of progression of motor complications. This pattern is consistent with the notion that lower dopamine transporter availability in SAA^−^ patients reflects greater loss of dopaminergic terminals, which reduces the capacity to buffer extracellular dopamine levels following levodopa administration^36^. This suggests that SAA status may reflect a dimension of dopaminergic sensitivity that amplifies both therapeutic response and susceptibility to treatment-related complications. A potential clinical implication of these findings is that SAA status may help inform treatment stratification and strategy in PD. For instance, patients with SAA^−^ PD may warrant a relatively more proactive and liberal symptomatic treatment approach, as they appear to show less sustained levodopa responsiveness over time and less prominent dyskinesias as the disease progresses.

Response to dopaminergic medication is common but not universal in PD, and reduced levodopa responsiveness can raise concern for atypical parkinsonism^37^. Although these patients showed longitudinal decrease in treatment responsiveness, this may be attributed to a faster decline in DAT availability. Furthermore, the presence of levodopa-induced dyskinesia, although not specific, is considered a supportive feature of PD diagnosis^38^, and its occurrence in other parkinsonian entities such as multiple system atrophy (MSA) follows distinct characteristics that may aid in clinical differentiation^39^. Interestingly, a meta-analysis showed that while striatal DAT function was reduced in both conditions, there was no statistically significant difference in overall striatal binding between PD and MSA-P^40^. Nevertheless, our study showed that the SAA^−^ sPD group had significantly lower dopaminergic reserve within their striatum. Together, these findings support the classification of SAA^−^ patients in our cohort within the PD spectrum and argue against the possibility that the observed differences are primarily driven by inclusion of atypical parkinsonism. That said, only continued clinical monitoring followed by postmortem validation can definitively establish their underlying PD diagnosis.

Sex-stratified analyses suggested that the association between SAA status and treatment response may be more pronounced in females, although these findings should be interpreted with caution. While female SAA^+^ patients showed a greater increase in medication responsiveness over time and a stronger divergence in ON-state progression, these effects were not consistently observed across all outcomes. Given the known sex differences in PD presentation and dopaminergic pharmacokinetics, these findings raise the possibility of sex-specific interactions between α-synuclein pathology and treatment response, but require replication in larger samples.

To the best of our knowledge, our study is the first report to compare levodopa responsiveness between SAA^+^ and SAA^-^ PD patients. These findings support the concept that SAA^−^ sPD represents a biologically distinct subgroup, potentially characterized by alternative or mixed pathophysiological processes. Recent studies have shown more prominent brain atrophy, overall rapid course of progression, and higher CSF pTau181/amyloid-β42 ratios, suggesting possible Alzheimer’s disease copathology as an alternative or contributing pathophysiology among individuals with SAA^-^ PD^41^. Future investigations are therefore warranted to elucidate the role and interplay of co-morbid pathologies with PD in SAA^−^ individuals.

Lack of external validation in independent cohorts of PD patients with SAA status available is a limitation of the present study. Longitudinal assessment of SAA status over time can also clarify dynamic changes and their relation to treatment response including any conversion from SAA^−^ to SAA^+^ status. We modeled treatment response as a linear trajectory; however, medication responsiveness may follow non-linear patterns over time. Future studies with larger sample sizes and visits per participant should explore non-linear dynamics in levodopa responsiveness across SAA groups. Nonetheless, to our knowledge, this is the first report directly comparing levodopa responsiveness between SAA^+^ and SAA^−^ PD patients.

In conclusion, CSF α-synuclein SAA status is associated with treatment-dependent differences in motor trajectories in PD. These findings suggest that SAA captures biologically relevant variability in responsiveness to dopaminergic treatment and may inform patient stratification and therapeutic optimisation.

## Data Availability

Data used in the preparation of this manuscript were obtained from the Parkinson's Progression Markers Initiative (PPMI) database (https://www.ppmi-info.org/access-data-specimens/download-data). All the information and data used in this study are publicly available for free and can be requested on the PPMI website (https://www.ppmi-info.org).

